# Genomic profile in TGCT Mexican patients reveals a potential biomarker of sensitivity to platinum-based therapy

**DOI:** 10.1101/2021.09.28.21264276

**Authors:** Rodrigo González-Barrios, Nicolás Alcaraz, Michel Montalvo-Casimiro, Alejandra Cervera, Paulina Munguia-Garza, Cristian Arriaga-Canon, Diego Hinojosa-Ugarte, Nora Sobrevilla-Moreno, Karla Torres-Arciga, Julia Mendoza-Perez, José Diaz-Chavez, Carlo Cesar Cortes, Ana Scavuzzo, Clementina Castro-Hernández, Jorge Martínez-Cedillo, Delia Pérez-Montiel, Miguel Jiménez-Ríos, Luis A. Herrera

## Abstract

Despite having a favorable response to platinum-based chemotherapies, ∼15% of Testicular Germ Cell Tumor (TGCT) patients are platinum resistant. Mortality rates among Latin American countries have remained constant over time, which makes the study of this population of particular interest. To gain insight into this phenomenon, we conducted whole-exome sequencing, microarray-based comparative genomic hybridization, and copy number analysis of 32 tumors from a Mexican cohort, of which 18 were platinum sensitive and 14 were platinum resistant. We incorporated analyses of mutational burden, driver mutations, SNV and CNV signatures. We observed that sensitivity to chemotherapy does not seem to be explained by any of the mutations detected. Instead, we uncovered CNVs, particularly amplification of 2q11.1 as a novel variant with chemosensitivity biomarker potential. DNA breakpoints in genes were also investigated and might represent an interesting research opportunity. Our data sheds light into understanding platinum resistance in a poorly characterized population.

## 1. Introduction

Testicular cancer accounts for approximately 1% of cancers in men, however, it is the most common tumor in males aged 15 to 44 (Znaor et al., 2014). Testicular germ cell tumors (TGCT) comprise 98% of all malignant neoplasms that arise in the testicle. Based on histological analysis TCGT can be classified as seminomas (SE-TGCT) and non-seminomas (NS-TGCT), both of which arise from germ cell neoplasia *in situ* (Ghazarian et al., 2017). Seminomas encompass 50-60% of the tumors, with a peak incidence at age 35. Non-seminomas comprise about 40-50% and are usually diagnosed around 25 years of age, the latter are noted for their diverse histological subtypes and varying degrees of differentiation (Gurney et al., 2019).

Since the introduction of platinum-based therapies, TGCT’s have been widely recognized for their outstanding 5-year survival rates. These values are close to 95% regardless of the histologic subtype and close to 80% for metastatic cancer (Allen et al., 2017). Nevertheless, platinum refractory disease - which is defined as persistent or rising tumor markers during or within 4 weeks of completion of a four-cycle platinum-based chemotherapy regime - is observed in approximately 10-20% of patients. Further therapeutic options are limited for these patients, and long-term survival rates remain poor. Understanding this phenomenon has been of broad interest to clinicians, but, to date, the mechanism leading to platinum resistance is poorly understood. In addition, upfront identification of platinum resistant or platinum sensitive patients using biomarkers is not feasible in the clinical setting, and the development of targeted therapies seems to be far-off (Jacobsen & Honecker, 2015).

Broadly, TGCT’s are characterized by frequent chromosomal anomalies, with polyploidization and gain of chromosome arm 12p as an isochromosome being the most regularly observed. Gain of chromosome X, 7, 21 and 22 are also frequent in these tumors (Litchfield et al., 2015). The mutation burden of this neoplasm has been described as low compared with that of other cancers; mutations in the *KIT* and *KRAS* genes have been highlighted as the most commonly reported driver genes for SE-TGCT (Shen et al., 2018).

To date, no mutations or genetic alterations have been described that could help identify resistance to platinum-based treatment. Several mechanisms such as defects in homologous recombination, *TP53* and *PTEN* mutations, higher mutational burdens and cell cycle regulation alterations, have been proposed as a possibility, however, none of them seem to adequately explain this phenomenon (Lobo et al., 2020).

Recently, efforts have been made to elucidate the genomic characteristics that underlie the TGCT subtypes. Whole Exome Sequencing (WES) has been one of the most widely used approaches. Through this method, some differences were noted between platinum resistant and platinum sensitive tumors. For instance, the mutational burden of resistant tumors seems to be higher than that of sensitive tumors. New putative driver genes and pathways like WNT/CTNNB1 have been associated with resistance and *KIT* and *TP53* mutations were also significantly different from one another (Loveday et al., 2020).

Incidence and especially mortality varies throughout different ethnicities, with non-hispanic whites being the most commonly affected population in the US. Nevertheless, hispanic population is the second most affected group (Li et al., 2020). Recently, chemoresistance has been broadly studied in american non-hispanic whites, caucasians and european populations since they tend to have the highest incidence (Loveday et al., 2020). However, chemoresistance in Hispanics has not been well studied. Although disease incidence seems to have stabilized, mortality has shown a different tendency. Unlike the US, Canada and Europe, where mortality rates have been decreasing, Africa, Asia, Latin America and the Caribbean’s mortality have remained stable (Park et al., 2018). These findings suggest that there’s an urgent need to explore these populations in order to improve early diagnosis and therapeutic approaches.

In this study, we carried out a genomic approach to broaden our understanding of TGCT platinum resistance in Hispanic populations by studying a Mexican cohort from the largest cancer center in Mexico using whole exome sequencing (WES) and microarray-based comparative genomic hybridization (aCGH) We observed a greater mutational load compared to that reported in other populations. We found mutated genes that are novel and could characterize our NS-TGCT patients. Also, we examined changes in copy number variants (CNVs) in greater detail and identified amplification of a novel region that could serve as a biomarker of sensitivity for platinum-based therapy. DNA breakpoints in genes were also explored, and interesting novel findings were revealed.

## 2. Methods

### 2.1 TGCT patient cohort

For this study, 32 patients with a clinicopathological diagnosis of type II or III Non-Seminoma TGCT were selected; the patients were treated between 2008 and 2018 at the Urologic Oncology Department at the National Cancer Institute of Mexico (INCan).

Patients underwent radical orchiectomy and were staged according to the American Joint Committee on Cancer (AJCC), 8th edition. Patients were classified according to the prognostic groups of the International Collaborative Group of Germ Cell Tumors (IGCCCG). All patients received treatment with bleomycin, etoposide, and cisplatin (BEP) as established by international guidelines. After treatment, the phenotypic response to chemotherapy (resistance or sensitivity) was evaluated. Platinum resistance was defined as disease that continually progressed under platinum-based chemotherapy, progressive disease (relapse or incomplete response) after one or more complete platinum-based regimes within the first 2 years of follow-up, viable (non-teratomatous) disease in a post-chemotherapy surgical sample, or persistent or rising tumor markers 4 weeks after a complete four-cycle chemotherapy regime (Loveday et al., 2020). Follow□up and re-classification were last updated in January 2021.

A summary of the study cohort is detailed in **Table 1**. The Ethics Committee and the Research Committee of INCan approved the study protocol **((012/031/ICI) (CEI/783/12)**. The research followed the tenets of the Helsinki Declaration based on approval by the hospital’s institutional review board and was conducted after obtaining the subject’s informed consent in accordance with institutional guidelines.

**Table 1.**
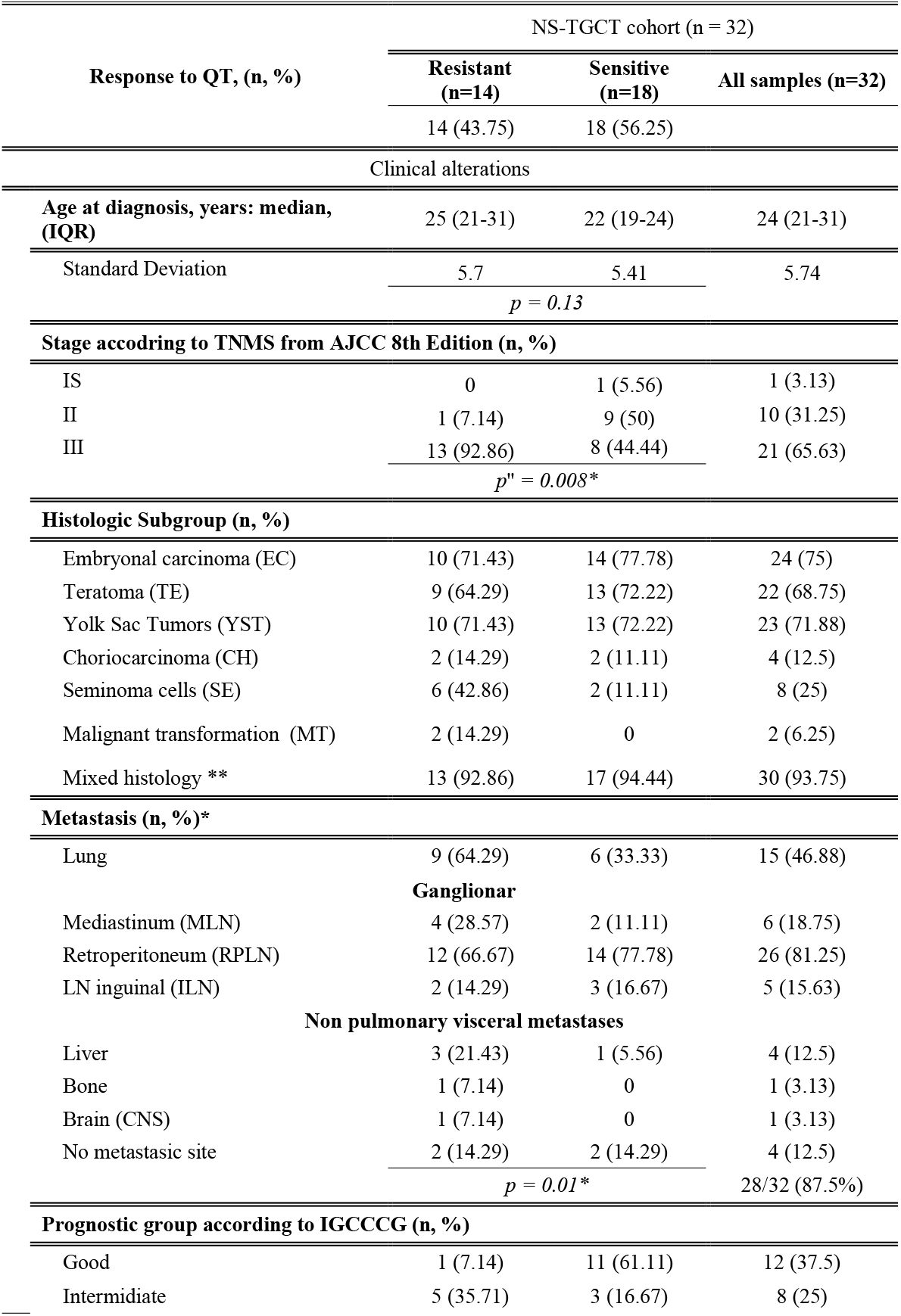
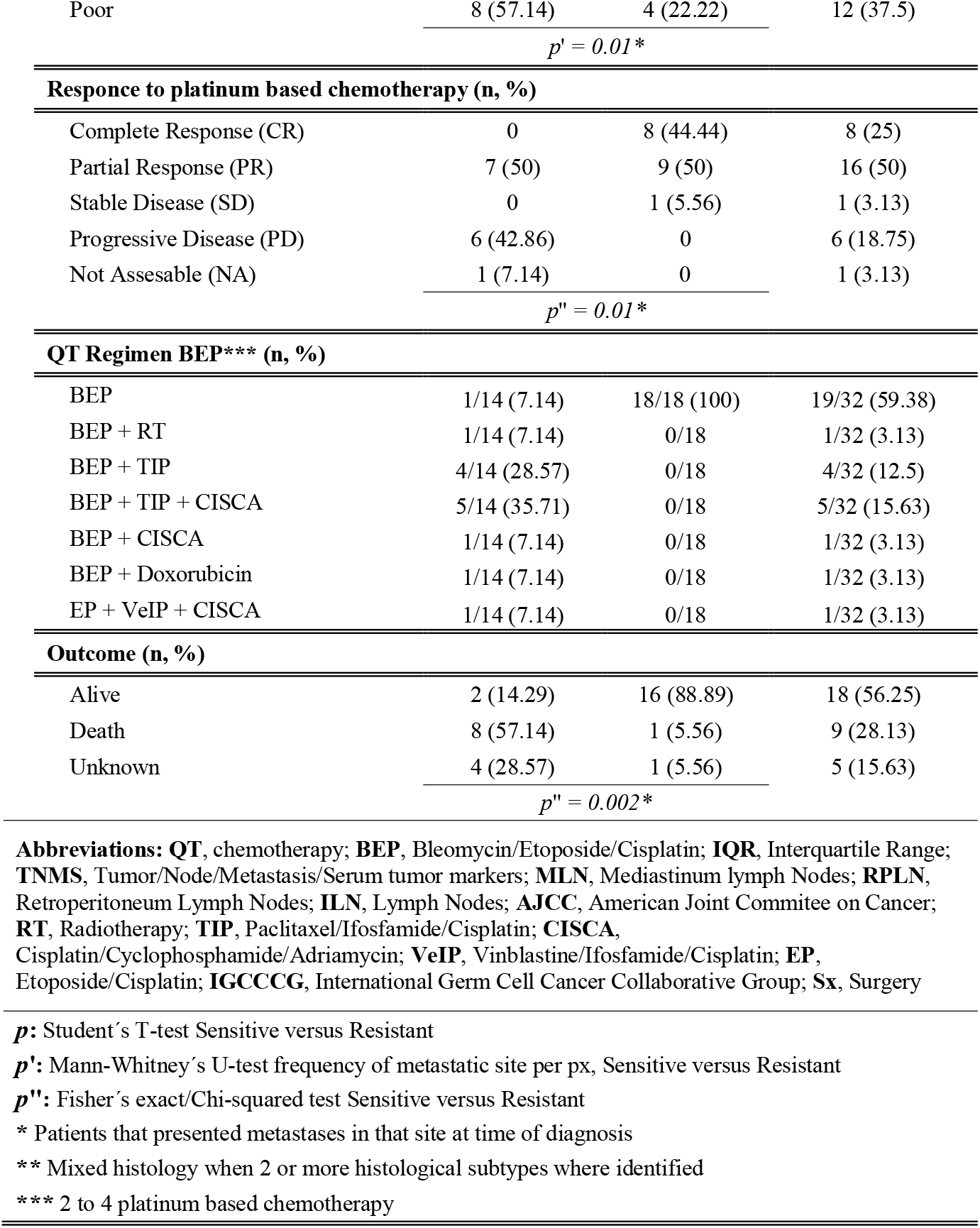
Clinical summary of Testicular Germ Cell Tumors Cohort

### 2.2 Sample collection and DNA extraction for WES

Tumor and peripheral blood samples were obtained at the time of radical orchiectomy of NS-TGCT patients. After the pathology department confirmed the histopathological diagnosis by evaluating the tissues, samples were frozen at -80°C 30 minutes after surgery (cold ischemia time) in RNA stabilization solution tubes. The tumoral tissues were routinely fixed in formalin and paraffin□embedded for subsequent histological examination. DNA was extracted from tumors and blood for sequencing using the DNeasy Blood & Tissue kit (Qiagen, Valencia, CA, USA) following the manufacturer’s specifications. The quality and integrity of the DNA was analyzed employing the TapeStation 2200 (Agilent technologies, Santa Clara, CA, USA) according to the specifications established by the manufacturer.

### 2.3 Whole Exome Sequencing (WES)

The complete exomes of 32 paired tumor tissue and peripheral blood (as control) samples were sequenced. From which 18 were platinum-sensitive and 14 platinum-resistant patients. Sequencing was performed using the HiSeq2500 Illumina platform, following the Illumina Nimblegen V3 protocol established at the Cancer Genomics Laboratory of the University of Texas MD Anderson Cancer Center (MDACC). The samples were sequenced at an average coverage of 100X for the tumor samples and 60X for the control samples.

### 2.4 Bioinformatics Analysis

#### 2.4.1 Preprocessing

Exome preprocessing was performed following GATK (v3.7) best practice guidelines. For Quality Control analysis of reads, we used FastQC v0.11.4. Reads were mapped to the human reference genome hg38 using BWA-MEM (version 0.7.12-r1039). Afterwards, duplicates were removed using picard (version 2.18.13). Base scores were recalibrated using the baseRecalibrator and ApplyBQSR tools within the GenomeAnalysis toolkit package to obtain a final set of analysis-ready bam files. Then, a Panel of Normals was assembled using the CreateSomaticPanelOfNormals (gatk-4.0.8.1) tool with all normal samples, which was subsequently used as input for somatic variant calling.

#### 2.4.2 Somatic variants (SNVs and indels) analysis

Somatic variants were called with Mutect2 (gatk-4.0.8.1) using tumor and control samples for each patient, the Panel of Normals, and the parameters minimum mapping quality of 50 and minimum base quality score of 26. Variants were filtered for contamination using FilterMutectCalls (gatk-4.0.8.1). Filtered variants were annotated and classified with Funcotator, (within gatk-4.0.8.1) tool. Analyses and plots of annotated filtered variants were performed using the maftools R package.

#### 2.4.3 Somatic copy number variations (SCNVs) analysis

For the detection, classification, and segmentation of SCNVs, the toolkit CNVkit (v.0.9.8) was used. Previously, the R package PureCN was employed to infer ploidy by purity of tumors, using this information to adjust the calls in CNVkit. To find significant recurrent CNVs in sensitive and resistant groups, GISTIC (v.2.0.22) and maftools R packages were used to visualize data.

### 2.5 Microarray based Comparative Genomic Hybridization (aCGH)

Array-CGH analysis was performed on 8 paired samples (4 platinum-resistant and 4 platinum-sensitive) from our TGCT cohort, using Agilent SurePrint G3 Human Genome microarray 4×180K (Agilent Technologies) targeting structural variations. DNA was extracted from frozen tumor tissue (see above) from TGCT patients whose platinum response was evaluated previously. For DNA extraction we used the DNeasy Blood & Tissue kit (Qiagen, Valencia, CA, USA).

The quality and integrity of the DNA were analyzed by TapeStation 2200 (Agilent technologies, Santa Clara, CA, USA) according to the specifications established by the manufacturer. Results were analyzed using Agilent Genomic Workbench (v6.5) and Agilent CytoGenomics (v5.1.2.1) software with the following settings: ADM-2 as aberration algorithm, threshold of 6 and moving average of 2Mb. The results are annotated according to Human Genome build 19 and include imbalances with at least three consecutive probes with abnormal log2 ratios.

### 2.6 Statistical analyses

The clinical variables of the patients included in this study were analyzed using descriptive statistics. Inferential comparisons were performed calculating the relative risk with their 95% confidence intervals. For the differences between groups, Student t-test or Mann-Whitney u-test was used depending on the nature of the data. Chi-Square and Fisher tests were used to compare the categorical variables. The alpha value was defined as p<0.05. All statistical tests were two-tailed.

### 2.6 Microarray based Comparative Genomic Hybridization (aCGH)

Array-CGH analysis was performed on 8 paired samples (4 platinum-resistant and 4 platinum-sensitive) from our TGCT cohort, using Agilent SurePrint G3 Human Genome microarray 4×180K (Agilent Technologies) targeting structural variations. DNA was extracted from frozen tumor tissue (see above) from TGCT patients whose platinum response was evaluated previously. For DNA extraction we used the DNeasy Blood & Tissue kit (Qiagen, Valencia, CA, USA).

The quality and integrity of the DNA were analyzed by TapeStation 2200 (Agilent technologies, Santa Clara, CA, USA) according to the specifications established by the manufacturer. Results were analyzed using Agilent Genomic Workbench (v6.5) and Agilent CytoGenomics (v5.1.2.1) software with the following settings: ADM-2 as aberration algorithm, threshold of 6 and moving average of 2Mb. The results are annotated according to Human Genome build 19 and include imbalances with at least three consecutive probes with abnormal log2 ratios.

## 3. Results

### 3.1 TCGT mutation burden is higher in Mexican patients, but does not define platinum resistance

We recruited a series of 32 non-seminoma TGCT cases comprising 14 platinum-resistant and 18 platinum-sensitive patients. (**Table 1**). Data from 21 metastatic and 11 primary tumors were evaluated and showed that most of our metastatic patients had more than one metastatic site, with non-pulmonary visceral metastases being fairly common. As expected, the majority of our patients had a mixed component histology, and pure tumors were rare among our cohort. We also included a pair of malignant transformations since they are quite unusual, and their particular resistance to chemotherapy has not been explored. All of our patients were classified according to their prognostic group, with a considerable proportion of them having poor prognosis from the beginning of their treatment. This information is relevant since progression-free survival rates are estimated based on this characteristic. Partial response was the most common outcome in our cohort which was expected since most of our patients had a teratomatous component in their tumor.

We extracted DNA from samples from each patient and performed WES assays, with a mean coverage of 100X across targeted bases. From the 32 cases a total of 1100 variants were called (**supplementary Fig 1**), in which missense mutations were the most frequent (800 variants). Single nucleotide variants (SNVs) were the most commonly found. We identified around 900 different SNPs present throughout all patients, these were mostly composed of C>A and C>T transversions (1105 and 1890 SNV classes respectively). As expected from previous reports, we observed a mean rate of 0.49 somatic mutations per Mb **(Table 2)**. This represents a low mutation burden, when compared to other solid tumours like melanoma and lung cancer, which present rates of 8.0 and 4.0 mutations per Mb, respectively. In this regard, we wanted to determine if the mutation burden of our patients was similar to that reported in the TCGA database. In general, we observed that our patients had a mutation burden rate greater than the average TGCT patients included, and such was the case even when we divided the patients into our therapy response groups. (**Fig 1a**). TCGT resistant and sensitive patients have mutation burdens around 0.53 and 0.43 mutations/Mb, respectively. Although we observed a trend for a greater mutation burden in therapy-resistant than sensitive patients, there was not a significant difference between both groups (**Fig 1b**). Therefore, our data suggest that there is a higher mutation rate in our population when compared to the one reported in TCGA (mainly Caucasian population); mutation rate does not impact the response to therapy based on BEP.

**Table 2.** Genomic alterations summary of NS-TGCT cohort

|  |  | NS-TGCT cohort (n = 32) |  |  |
| --- | --- | --- | --- | --- |
| Response to QT (n, %) |  | Resistant<br>(n=14) | Sensitive<br>(n=18) | All samples<br>(n=32) |
|  |  | 14 (43.75) | 18 (56.25) | 24 (16-36) |
| Genomic alterations |  |  |  |  |
| Mutation rate/Mutations per Mb, median, (IQR) |  |  |  |  |
| Somatic<br>Variants<br>(SNV) | Tumor Mutation Burden<br>(TMB) | 0.53 (0.3-0.6) | 0.46 (0.3-0.5) | 0.49 (0.3-0.6) |
|  | Standard Deviation | 0.19 | 0.21 | 0.20 |
| | $p = 0.36$ | | | |
| Tumor Ploidy & Purity, mean, (IQR) |  |  |  |  |
| Copy<br>Number<br>variations<br>(CNV) | Ploidy (Absolute) | 2.60 (2.6-4.0) | 2.84 (2.1-3.0) | 2.92 (2.4-3.3) |
|  | Standard Deviation | 0.98 | 0.57 | 0.77 |
| | $p' = 0.17$ | | | |
|  | Purity (Absolute) | 0.54 (0.3-0.6) | 0.44 (0.2-0.6) | 0.48 (0.3-0.6) |
|  | Standard Deviation | 0.21 | 0.16 | 0.75 |
| | $p = 0.13$ | | | |

$p$ : Student's T-test Sensitive versus Resistant
$p'$ : Mann-Whitney's U-test Sensitive versus Resistant

**Fig 1.**
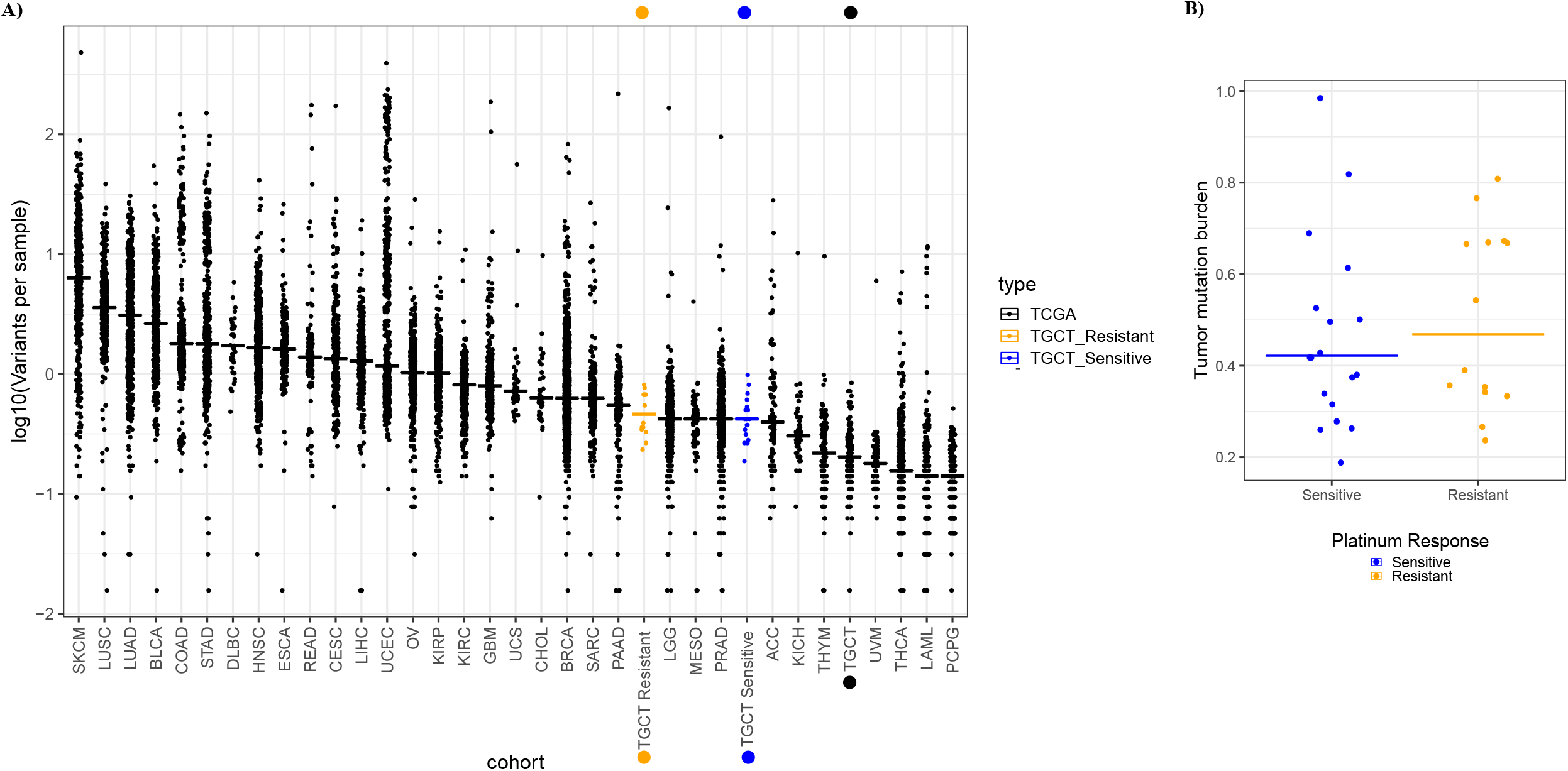
Comparison of tumor mutation burden in INCan-TCGT cohort vs TCGA. **A)** Median frequencies of somatic variants reported in exome sequencing (horizontal lines) across multiple tumor types reported in TCGA. Left to right, highest and lower frequencies, as measured in mutations per megabase (Mb). Broadly, the INCan-TCGT cohort (orange and blue dots) has shown (both platinum sensitive and resistant samples) that mutation rate is higher than reported in TCGA for TGCT tumors. **B)** Discrimination of mutation burden by response. Platinum-resistant samples (orange dots) have shown higher (yet not significant) frequency in variants per sample in comparison with platinum-sensitiveTGCT samples.

### 3.2 Gene variants are not sufficient to characterize response to therapy

To understand possible driver mutations, or those that could serve to identify therapeutic responses, we first explored the general features of our patients and their association with the clinical outcome, shown as clinical response and prognostic groups. As described in **Table 1**, we observed that our sensitive and resistant patients presented a heterogeneous histological subgroup, most of which was represented by the mixed type. Moreover, most of the resistant patients were stage III and classified into the poor prognosis group. When analyzing their clinical response, we found either progressive or stable disease. Whilst sensitive patients who, as expected, had a good prognosis, 44% presented a complete response to therapy. Interestingly, the mutation burden was not directly associated with the prognostic group. Regardless of the mutational burden, patients were still classified into the poor prognostic group, suggesting that mutation accumulation does not drive disease outcomes or the response to platinum-based therapy (**Fig 2a)**. Next, we performed an analysis of the genes most significantly altered in all the patient samples based on their responses. Although we found *KRAS* mutations in 6% of the patients, this was not the most frequent mutation. In our samples, mutations affecting *COL6A3, NCOA3, TNR* and *ZFHX3* were the most abundant (with 9% frequency each). In most of the genes, missense mutations were the most frequent type of mutation, however, some mutated genes such as *NCOA3, ZFHX3 and ANGEl1*, presented other varieties of mutations (**Fig 2b)**. Taken together, these findings reveal that, except for the variants mentioned, most of the mutations seem to be random and infrequent in the different response groups. This suggests that mutations are not the direct cause of resistance to therapy, but they could lead to the risk of developing these types of tumors.

**Fig 2.**
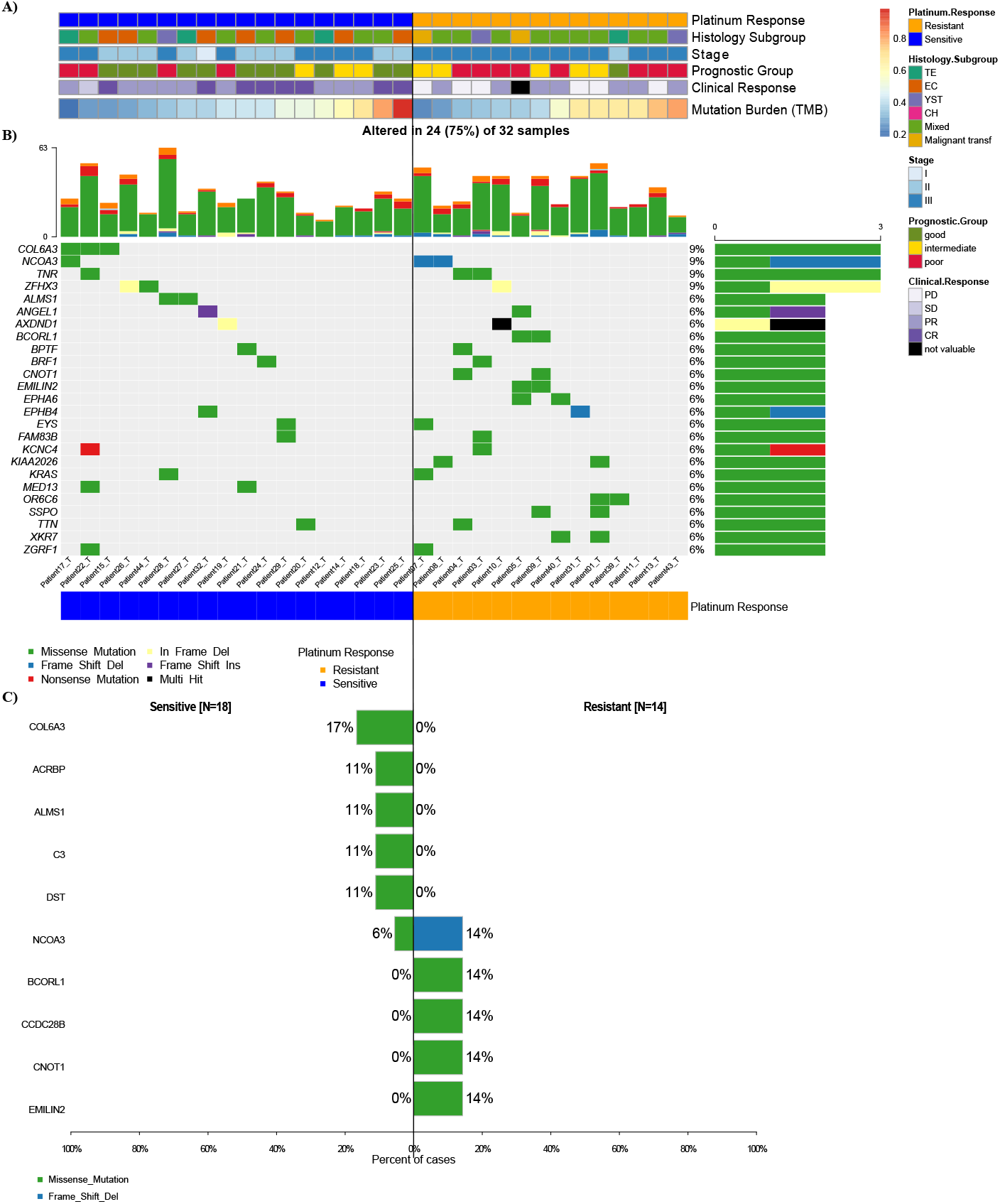
Mutational landscape of cancer driver genes in TGCT. **A)** Heatmap with clinical features of patients, each row represents a sample divided by response group and another clinical characteristics **B)** Oncoplot showing the most frequently mutated coding genes across 32 NS-TGCTs by response groups. Each column represents a sample and each row a different gene with associated color for each mutation type. The top bar plot has the frequency and type of mutations for each patient, while the right barplot has the frequency of mutations in each gene. Samples are ordered by the platinum response group, resistant and sensitive (orange and blue, respectively). The highest frequency of mutations in each gene is 9% and represents all random mutations observed in at least two samples. **C)** Most frequently mutated genes and percentage of cases where these mutations were found separated among groups, sensitive (left) and resistant (right). Neither genes are significant in this discrimination.

Our results show that the variants identified do not seem to provide a clear pattern that can be used to classify patients in terms of response to therapy, but rather appear to be random. Furthermore, statistical analysis using forest plots indicated that there were no significant differences between our groups (**Fig 2c**). Nonetheless, we managed to detect mutations in genes such as *COL6A3, ACBRBP, ALMS1, C3* and *DST* (with a frequency between 17 and 11%) that are exclusively seen in sensitive patients. We also observed variants unique to platinum-resistant patients, such as *BCORL1, CCDC28B, CNOT1* and *EMILIN2* (observed with a frequency of 14% each) (**Fig 2c**). These findings do not detract from the fact that these variants might possess biological significance. Of all the most frequent mutations, the *COL6A3* gene (9% frequency in NS-TGCT tumors) is the most representative one. Although this mutation is present in all sensitive patients, it is not suggested to be a significant sensitivity biomarker, but rather to be relevant as a driver gene for testicular cancer, nonetheless, a larger cohort is required to validate this result.

Taken together, our results show, as expected, that platinum-resistant patients have a worse prognosis and clinical outcome. Our data suggest that the observed mutational variants are not capable of defining response to platinum-based therapy. However, we identified different genes that could behave as potential drivers for NS-TGCT disease.

Since individual genetic variants do not define response to therapy, we wanted to assess if the phenomenon was linked to specific cell pathways that could jointly distinguish sensitivity or resistance to treatment. We observed that there were pathways that were affected in more than 50% of patients, among which the most affected were RTK-RAS, NOTCH, WNT, HIPPO and PI3K. Specifically, the MYC pathway was more affected in resistant patients, while the NRF2 pathway was only identified in sensitive patients. However, as a whole, the affected pathways are independent of the response to platinum-based treatment. Thus, our results suggest that these pathways are tumor-dependent or specifically more affected in NS-TGCT patients, which could therefore suggest an indicator of risk for developing the illness (**Fig 3a and b**).

**Fig 3.**
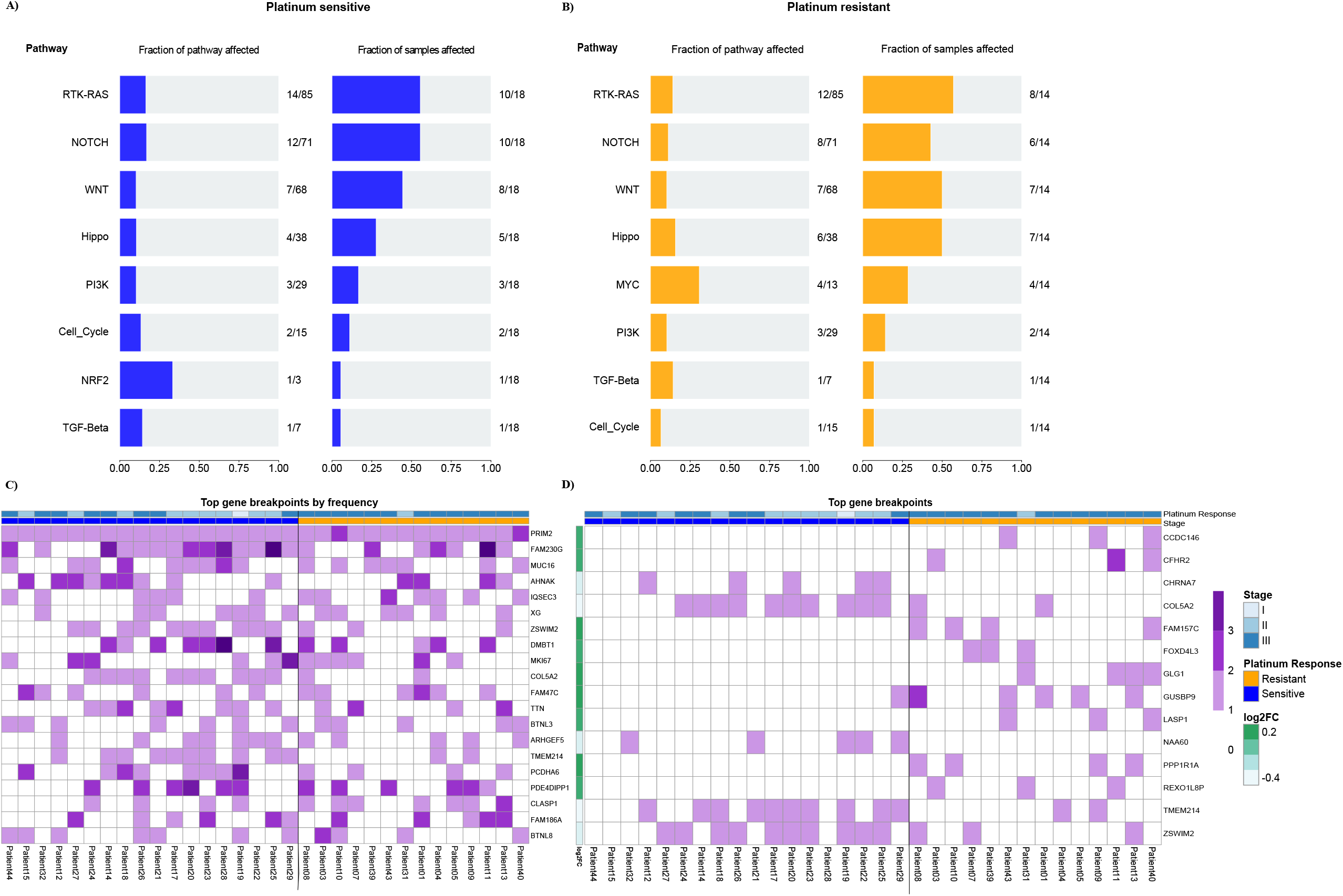
Oncopathways and genomic breakpoints analysis. **A) & B)** Frequency plot of oncopathways affected in platinum-sensitive and platinum-resistant samples, shown left to right, fraction of the pathway affected and the fraction of samples that present an event located in these pathways. There are not the same oncopathways affected between the two response groups. **C)** Gene Breakpoint analysis, derived from CNVKIT: Top 20 most frequent genes of all the genes that had breakpoints in the whole cohort **D)** Oncoplot with most frequent breakpoints in genes that greatly separated the two conditions. The fold change is the log-ratio of the fraction of patients between resistant and sensitive that have those breakpoints. Log2 (fraction of breakpoints in resistant) / (fraction of breakpoints in sensitive)

### 3.3 DNA breakdown sites are frequent and differ in groups of patients with different responses to platinum-based therapy

It is known that chromosomal instability, especially DNA breaks, are of special relevance in cancer. We evaluated the frequency of genetic breakpoints that occurred in our patients. We observed that in most of our patients, a large number of these were concentrated in specific genes. This is the case for those present in *PRIM2, FAM230G* and *MUC16* genes. This result seems relevant to us since the mutations that we observed previously are random in nature, but the breakpoints are not. In fact, these breakpoints were demonstrated to be consistent hot spots in our patients. Some of these breakpoints have more than 1 cleavage site, such as those in the *FAM230G* and *AHNAK* genes (**Fig 3c**). Interestingly, all the patients presented breakpoints in the *PRIM2* gene. *PRIM 2* is a regulatory subunit of the DNA primase complex and component of the DNA polymerase alpha complex, which is highly relevant for the initiation of DNA synthesis. Likewise, breaks in the *MUC16* gene are present in 50% of patients. *MUC16* is overexpressed in multiple cancers and plays an important role in tumorigenicity and acquired resistance to therapy. *MUC16* (usually referred to as *CA125*) is widely known and has been extensively used as a biomarker for ovarian cancer, and its expression has been associated with disease progression.

Then, we wanted to determine if there were specific breaks among the clinical groups and their response to therapy. Therefore, we filtered them based on the log2 ratio between the frequency of breakpoints in the resistant cohort versus the sensitive one. We observed that breakpoints were more frequent in resistant patients, where the most constant breakpoints were those that occurred in *CCDC146, CFHR2FAM157C, GUSBP9* and *PPP1R1A* genes. In therapy-sensitive patients, we found less frequent rupture sites (6 genes) present in the *CHRNA7, COL5A2, NAA60, TMEM214* and *ZSWIM2* genes (**Fig 3d**). These results suggest that DNA breaks occur at common sites in patients and that these breakpoints could be related to the phenomenon of chemoresistance. Therefore, this result suggests a panel of ruptures that are able to cluster and distinguish the response to platinum-based therapy in NS-TGCT patients, especially in Mexican populations.

### 3.4 Copy Number Variation analysis could define differential molecular signatures for platinum sensitivity

Copy number analysis was performed on the 32 primary TGCTs using WES, with complete clinical data available (**Table 1)**. The absolute mean tumor purity was >40%. There is heterogeneity among the samples and a tendency for a higher purity on chemoresistant samples than chemosensitive samples. (median= 0.54 y 0.44 respectively). The majority of tumors showed hyperploidy (72% had ploidy >2.5) **(Table 2)**. With this data, the calls in the CNVkit tool for segmentation and detection of significant CNVs were adjusted **(Fig S2)**.. Broadly, TGCTs exhibited an elevated rate of arm-level amplifications (median number of arms/tumors with ≥1 allele amplified = 9). The most frequently observed individual arm-level events included gains of 12p (90%), 20q (62%), 2p (76%), 7p and 8p (74%). The most frequent focal events were amplification of 12p13.33 (96%), 12p12.1 (93%), and 12p11.21 (92%) **(Fig 4a)** as well as deletion of 4q and 5q (67%), 10q and 11q (65%) 13p, 18q and Y (90%). Additionally, we found around 29 amplification sites and 52 deletion sites (arm-level) distributed throughout the genome in all samples **(Fig S3)**.

**Fig 4.**
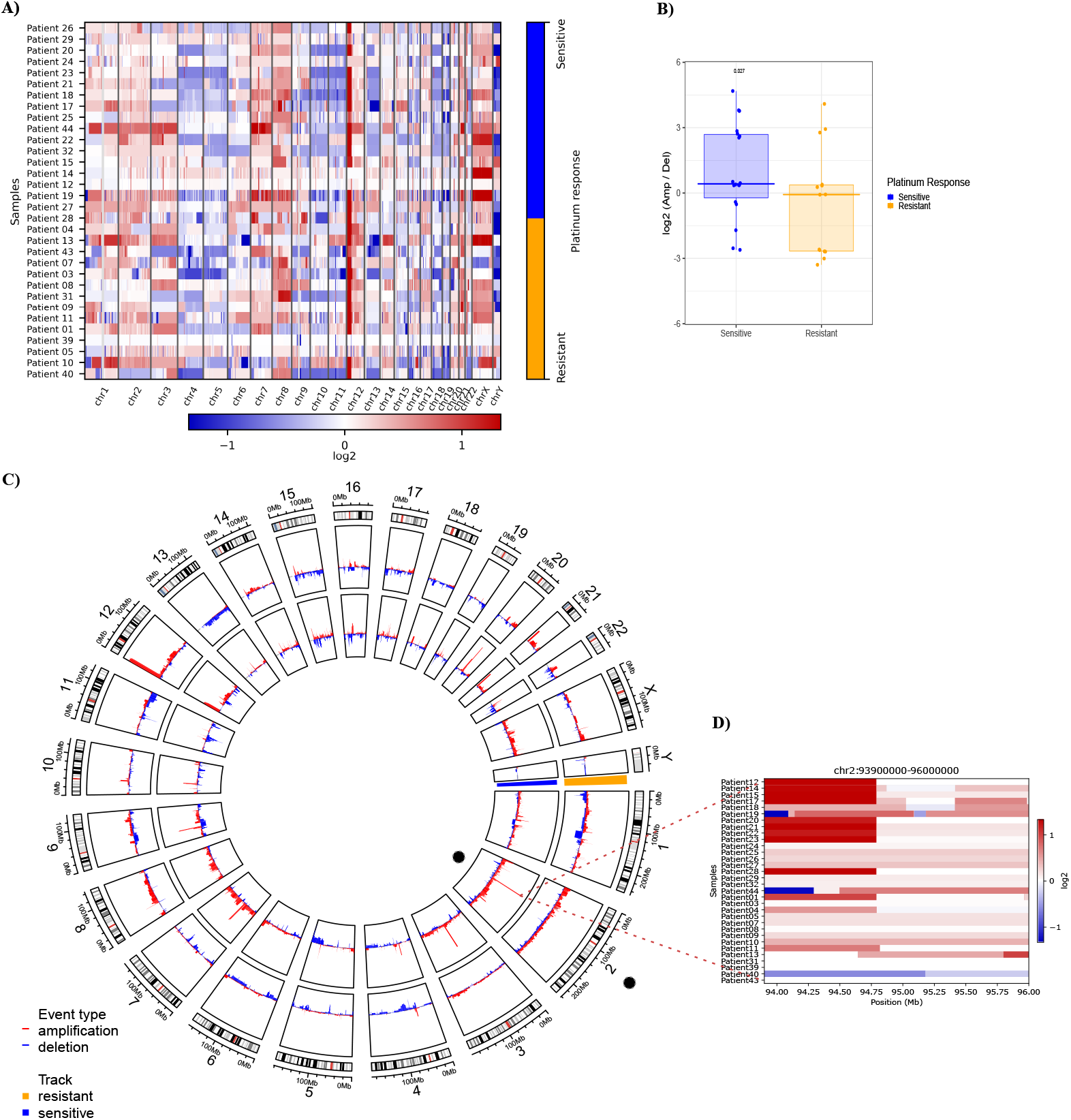
Significant CNVs segments that can distinguish between resistant and sensitive response to platinum. **A)** Heatmap of calls in copy number variations (either bins or regions). It shows an overview of the larger-scale CNVs for all our samples (both platinum responses), bins of gain (red) and loss (blue) are in color range for all genome arrays showing low and higher-amplitude segments. **B)** Heatmap from CNVKIT focused on chr2, each row represents a patient sample and each column represents an event (gain in red, loss in blue) that can occur in this position (Mb). Zoom shows a heatmap focused on events in the 2q11.1 region, Gain events are mostly common in sensitive samples. **C)** Circos plot of arm-level events obtained with GISTIC analysis, separating sensitive (inner circle, blue) from resistant (outter circle, orange) **D)** Boxplot of log-ratio of amplifications over the losses, we see that the sensitive samples tend to have this ratio higher than the resistant ones (analysis exclude X, Y chromosomes, p-value from wilcoxon test).

To determine whether there were differences between the amplification/deletion events present in platinum-sensitive and platinum-resistant patients, we performed a Wilcoxon test on autosomal chromosomes of all samples, with the log-ratio of amplifications over losses. We observed that platinum-sensitive samples tend to have a significantly *(p=0.027)* higher ratio than platinum-resistant samples. 12p, 2q, 3p, 7q were the most frequent amplifications. This suggests that platinum-sensitive samples have more amp/del events than resistant samples **(Fig 4b)**.

Aiming to evaluate whether the differences between platinum-sensitive and platinum-resistant patients are focused on arm-level events, we made a multifactorial adjusted analysis on the frequencies of global, individual arm or focal chromosomal aberrations **(Fig 4c**). We found important differences in focal CNV variation sites among both response groups, reported as false discovery rate corrected (*G-Score* value), estimated for genomic segments.

In order to assess if any of these alterations in CNVs was able to distinguish chemoresponse, we performed linear modeling with limma on the actual changes in copy number values as defined by GISTIC to identify significant gains or losses in arm bands between the resistant and sensitive groups. We found 2q11.1 to be the only significant aberration (adjusted p value= 0.03) **(Fig 4c)**. Figure 4d shows a heatmap of the chromosome 2 genomic region and the q.11.1 sub-arm region **(Fig 4d)**, which highlights the difference between the frequency of gains among sensitive and resistant samples. Our results suggest that a higher number of gain events in the region is strongly correlated with platinum sensitivity. Therefore, we suggest that focal gains in segment q11.1 of chr2 could be a biomarker of sensitivity to platinum-based therapy.

We found important differences in focal CNV variation sites among both response groups, reported as false discovery rate corrected (G-Score value), estimated for genomic segments. Broadly, G-Scores values by frequent segments indicate that resistant samples show less variation in terms of broad events **(Fig S3**), than sensitive ones. Platinum-sensitive samples show a greater number of events for both gains and losses, accentuating a higher density in gain events in regions where platinum-resistant remained unchanged.

Moreover, platinum-resistant tumors (**Fig 4c)** showed an increased frequency of gains in 12p13.31 compared to platinum-sensitive tumors. Also, our results show certain regions with different gain or loss events between sensitive and resistant tumors, but for the most part, events in various segments are repeated between both response groups **(Fig S3)**.

To test this model, we evaluated possible differences between the response groups through the most aberrant copy number analysis. We determined the top 20 recurrent arm-level CNVs according to the GISTIC analysis for only resistant samples **(Fig 5a)** and for sensitive samples **(Fig 5b)**. The results show an important difference between the amplification and deletion events. Platinum-sensitive samples had a greater number of amplification events than platinum-resistant ones, which seem to have mostly deletion events. In resistant samples, the most frequent amplification segment was 12p13.31 (86%) **(Fig 5a)**, whereas in sensitive samples, amplification of 8q12.1 was present in 100% of patients **(Fig 5b)**. Furthermore, it is important to note that the difference between the amp/del events between both response groups does not seem to be conditioned to late stages of cancer, since in both, early and advanced stages, the frequency of amp/del seems to stay the same.

**Fig 5.**
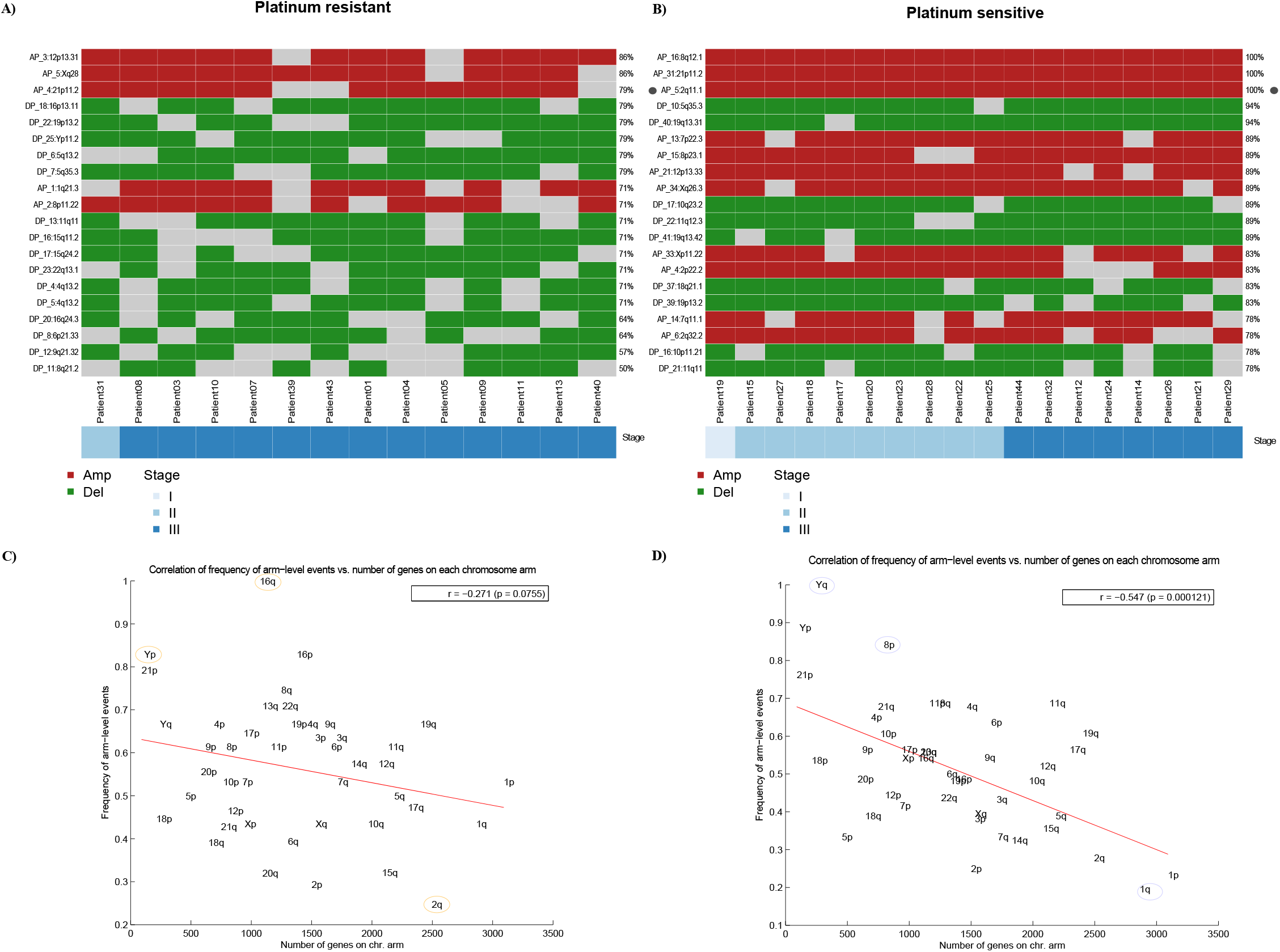
Most frequent arm-level amplifications/deletions events between sensitive and resistant TGCT. **A) & B)** Heatmap of arm-level regions top 20 in platinum resistant samples, each row represents a segment with its frequency, and each column a patient of the cohort that presents an event in these regions. Ordered by stage. Left to the right, resistant and sensitive **C) & D)** Correlation (with significant p value) between frequency of arm level events (Y) and number of genes in chromosome arm (X) in platinum resistant and platinum sensitive respectively. Anticorrelation (-r) is higher in sensitive samples, left to the right, resistant and sensitive.

To evaluate whether the most frequent loss or gain events are limited to areas where there is a higher density of genes, we performed an anti-correlation analysis that showed that a higher density of genes implies fewer loss/gain events **(Fig 5)**. Resistant samples tend to have higher losses/gains in gene-dense areas **(Fig 5c)** than do platinum-sensitive. Conversely, the anti-correlation is more pronounced in platinum-sensitive samples (**Fig 5d)**, which reveals that they tend to have a greater number of loss/gain events in areas of lower gene density than do platinum-resistant.

### 3.5 Array-CGH validates copy number variations on genomic regions finding in WES

According to previous exome studies, the gold standard for the validation of copy number variation data in WES analysis is CGH array. Therefore, to confirm our previous results, we performed a genomic hybridization microarray. Comparison of Agilent SurePrint G3 with 4×180K hybridization probes focused on the search for CNVs, in 4 platinum-sensitive tumors and 4 platinum-resistant tumors paired with reference DNA from each individual belonging to the same cohort sequenced by WES **(Fig 6)**. CNV call profiles were constructed individually for each sample, finding sites with more informative CNVs for each probe on all chromosomal arrays. CGH array also showed a variation in CNV significant calls per sample in most resistant cases, which were reported as >50 (i.e., there are >50 bins or regions with CNVs events, either Amp, Del, Gain or Loss). These results should be able to distinguish between the two platinum response groups and could define molecular signatures based on frequent CNVs. The concordance for CNVs between the exome analysis and the aCGH was 80% according to the average of amplifications and deletions, as was the concordance between the focal events compared with the segmentation of the GISTIC and CNVKIT tools used in the WES analysis.

**Fig 6.**
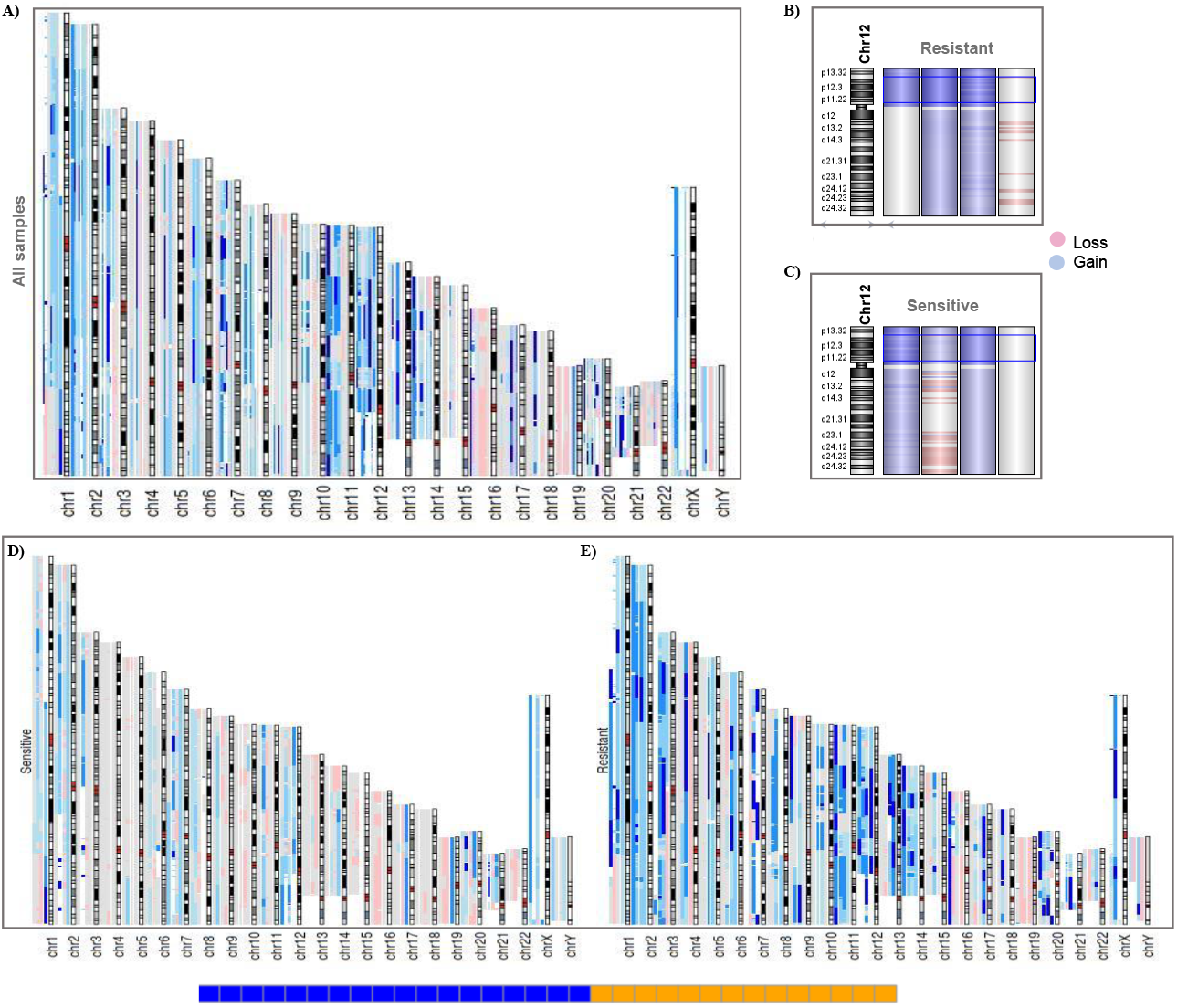
CGH array reveals a genomic landscape of instability in TGCT patients. **A)** aCGH of all samples, each sample with large events in the chromosome array was aligned in the same position. (gain in blue, loss in red) deep colour in a region represents a higher frequency of incidence (two or more samples) of events in the region. **B) & C)** Resistant (up) and Sensitive (down) samples in chromosome 12, showing that sensitive and resistant ones had 12p amplification, except in a case of each group. **D) & E)** Resistant and Sensitive ones aligned separately to view the differences on frequency of gain and loss events, shown that resistant ones present more events of gain than sensitive samples.

As expected, the aCGH revealed a general landscape of genomic instability with more frequent events of gains rather than losses **(Fig 6a)**. Broadly, all variation sites shown in WES (GISTIC analysis) were also found in aCGH due to genomic hybridization deep recovery probes used (4×180K). For instance, gains of chrX and losses of chrY were consistent in all samples (both previously reported in literature). This did not occur with isochromosome 12p amplification, which was not found in a sensitive sample. However, these results were consistent with the WES analysis, since both samples did not show the i12p amplification **(Fig 6b and c)**.

Also, aCGH revealed a general landscape in which resistant samples present more amplification events compared to sensitive samples **(Fig 6d and e)**. This suggests that resistant patients had higher genomic instability than platinum-sensitive tumors. Finally, we show chromosome 2 and the amplified segment 2q11.1 in a platinum-resistant sample **(Fig S4a)** and a platinum-sensitive **(Fig S4b)** sample to compare with the relationship found previously, where the gain events in the region seem to define the innate sensitivity of the patients. The occurrence of this event is shown by an opening in the region, where the genes that make up the region are laid out with their respective gain events.

## 4. Discusión

Testicular cancer is known to have a low incidence when compared to other neoplasms, however, its incidence is quite variable when comparing different countries. For example, when incidence is analyzed in Mexico, whilst it is expected to account for 5% of urological cancers (Vasdev et al., 2013), the latest report from central Mexico (where our oncologic center is held) stated a proportion of 25%, which is just below prostate cancer (Jiménez-Ríos et al., 2011). Additionally, mortality rates have always been low and have been stabilizing or decreasing worldwide. This has been attributed to better screening techniques, prompt diagnosis and the widely recognized sensitivity to platinum-based chemotherapy. However, for Mexican populations, mortality rates have remained unchanged, which might suggest that platinum-resistance is fairly common among our population. To date, most of the genomic studies of TGCT have focused on characterizing the disease specifically risk factors, and these have not focused on elucidating resistance to chemotherapy, which remains an unresolved clinical issue (Rapley et al., 2009; Chung et al., 2013; Batool et al., 2019). Previous genomic studies have aimed to unravel this phenomenon, nonetheless, they have been focused on Caucasian populations. Hispanic populations have not been brought upon and remain to be broadly understudied. To our knowledge, we have assembled the largest NS-TCGT WES series from Hispanic-Mexican patients to date. We identified a series of novel mutated genes that have not been described (Litchfield et al., 2015; Shen et al., 2018; Loveday et al., 2020).

In concordance with previous studies, we observed that NS-TGCT patients have a low mutation rate compared to other tumors (Shen et al., 2018; Litchfield et al., 2015). Unexpectedly, our patients had a higher rate than previously reported for TGCTs and according to earlier reports this rate was slightly higher in patients resistant to platinum-based therapy than those sensitive (Bagrodia et al., 2016). Interestingly, although we detected mutations in *KRAS*, these were not the most abundant in our cohort. A frequency of 6% was observed when analyzing *KRAS*, differing from results in other populations where it has been as high as 12% (Necchi et al., 2018; Shen et al., 2018; Loveday et al., 2020). When compared to other studies mainly focused on Caucasian populations, the most frequent set of mutations that we found, headed by the *COL6A3, NCOA3 and TNR* genes, are not usually reported in the top 20 variants. Additionally, we did not find mutations in the *KIT* or *TP53* genes, which have been described as frequent in TGCT. A recent study observed that although *KIT* has frequent mutations, they are mainly present in SE-TGCT tumors which explains why it was absent since our population was mainly NS-TGCT (Necchi et al., 2018; Litchfield et al., 2014). Taken together, our data suggest that both the mutational burden and the mutational profile are different in patients of Mexican origin.

Some studies refer to mutations in *TP53* as well as alterations in *MDM2*, as characteristic of platinum-based resistance phenotype and an indicator of poor prognosis (Necchi et al., 2018). This was not the case in our study, since the variants were not among our results. Although the variants were not significantly able to clearly define resistance or sensitivity to platinum, we detected those that showed a tendency to be specific for sensitivity (*COLA6, ACBRBP, ALMS1, C3* and *DST)*, with frequencies between 17 and 11% in these tumors, and those with a tendency toward platinum resistance *(BCORL1, CCDC28B, CNOT1* and *EMILIN2*) that presented a frequency of 14%. We suggest that they may be considered potential biomarkers that require a larger study for their validation and future use in the clinical setting.

Interestingly, we revealed that individual genetic variants do not appear to define the response to platinum-based therapy in NS-TGCT patients. However, variants frequently occur in 8 main molecular pathways, headed by RTK-RAS, NOTCH and WNT, these have at least 1 to 2 mutated genes present in the pathway. Although they do not define resistance, it seems that they are the main pathways involved in the development of NS-TGCT in our population. In contrast, studies focused mainly in Caucasian populations found RAS-RAF, PI3K/MTOR and WNT/CTNNB1 as the mainly affected pathways which highlights the need to study Hispanic populations (Loveday et al., 2020; Shen et al., 2018). Nevertheless, more studies will be required to understand the importance of these variants as risk biomarkers for the disease.

It is known that these tumors present clearer indicators of numerical and structural variations. We addressed the possibility that the presence of DNA breakpoints could be relevant in the etiology of the tumor and in the response to platinum-based therapy (Hoff et al., 2016). Remarkably, we observed the presence of DNA breakpoints that were characteristic of NS-TGCT independently of the response to therapy and those that could serve as a panel to distinguish between patients from the sensitive and resistant cohort. To our knowledge, this is the first time that this has been reported and observed in the Hispanic population. The relevance of these breakpoints for their use in the clinical setting is a new field to explore in future research.

Structural and CNV variations have recently gained importance in the setting of describing TGCTs. As previously described, we found that TGCTs are mostly characterized by structural aberrations since the development of the disease, probably due to early non-disjunction of Primordial germ cells that lead to amplifications and deletions of both focal and arm-level chromosomal regions like the 12p isochromosome (Bryce et al., 2019). Overall, we found that the high frequency of large-arm CNV events in NS-TGCTs does seem to be different among sensitive and resistant patients in accordance with previous studies. (Loveday et al., 2020) However, CNVs analysis revealed that genomic instability in sensitive patients is slightly higher than in resistant patients with a clear pattern in some focal amplifications with high copy number states as gains in 8q12.1 and 21p11.2. Particularly, gains of chromosome arm 2q11.1 is present in 100% of sensitive NS-TGCT, and it was significant with chemosensitivity. Recurrent gains of 2q11 have not been observed in TGCT but it has been previously associated with neurological disorders (Riley et al., 2015; Kresse et al., 2008), thus, it could be considered as a novel biomarker of sensitivity to platinum chemotherapy. Rather than a mere passenger effect, the focal amplification in sensitive patients may reflect an active alternative mechanism by which tumor cell death is promoted and sensitize tumor cells to therapy (Vékony et al., 2009; Jeong Kwon et al., 2017).

Finally, our findings are consistent with previous works focused on TGCT-CNVs analysis. However, we found a higher frequency of gain or loss events in the sensitive patients, than the resistant ones, which is discordant to other populations studied (Loveday et al., 2020). Such discordances between sensitive and resistant patients could be due to variations in the ploidy, and tumor purity of our cohort. Therefore, with the aCGH validation, even the focal events with the lowest frequency in CNVs barely detected by WES (which could not be informative by themselves), helped us to confirm that CNVs and structural variations are more common in sensitive patients. Notwithstanding our principal limitation is the size of our cohort due to the low frequency of NS-TGCTs platinum-resistant patient availability, in this regard, another study with more samples, also suggested predominance of molecular signatures with inverse association with platinum resistance (Loveday et al., 2020). This suggests that response to platinum-based chemotherapy is of multifactorial origin and further studies are required.

Taken together, our results show an advance in the characterization and genomic exploration of NS-TGCT in a population of Latin origin. Suggesting that even with similar methodological processes, there’s a heterogeneity among genome of different populations, which strengthens the need to explore more largest Latin populations.

## 5. Conclusion

Our results showed that the NS-TGCT cohort of Mexican patients analyzed by WES presented a higher mutational burden than what has been reported internationally in TCGA for testicular germ cell tumors. The mutation burden in platinum-resistant patients shows a tendency to be higher than the one of platinum-sensitive patients. However, we did not find high-frequency mutated genes that are related to oncopathways or DNA-repair. Therefore, we suggest that in our cohort, the acquisition of resistance is not linked to an increase in mutations on oncodriver genes. In addition, we found a high frequency of breakpoints in relevant genes throughout the entire cohort, some of which could serve as a potential panel to differentiate between the two platinum response groups. They could also define the principle of genomic instability or the etiology in NS-TGCTs.

Our data revealed that our cohort presents genomic instability, characterized by a wide list of altered segments throughout the genome. Moreover, higher CNV events were found in platinum-sensitive patients than platinum-resistant ones. Finally, we found that gains in the 2q11.1 segment significantly (adjusted *p value= 0.03*) distinguished platinum-sensitive from platinum-resistant patients; therefore, this could be considered as a potential biomarker of sensitivity to platinum-based chemotherapy in Latin origin patients.

## Data Availability

Are currently being uploaded to GEO database.

## Acknowledgements

This work was supported by a grant CONACYT **(RG-B, 2900441)**. We thank Gaudencio Reyes-Vargas, MD., and Miguel Angel Jiménez-Dávila, MD., for his help collecting surgical tumor tissues and obtaining informed consent for each patient. We thank Diego Díaz-García. MD., for his help in staging and re-classification of the INCan-TGCT cohort.

PMG submitted this project as a research intern from the Dirección General de Calidad y Educación en Salud, Secretaría de Salud, México.

## Figure legends

**S1**.

Summary of coding somatic variants found on NS-TGCT cohort **A)** Frequency and classification of variants found (missense). **B)** Types of mutational variants most frequently found (SNP). **C)** Transversions and transitions represented in the cohort **D)** Variants found per sample (median 27 variants). **E)** Distribution of variant types found per sample (color scheme is the same as **A). F)** Top 10 genes mutated on the cohort and mutation types.

**S2**.

Comparison of tumor ploidy and purity median from the sequenced samples obtained by PureCN analyses. **A)** Tumoral purity reported for chemoresistant (left, orange) and chemosensitive (right, blue) NS-TGCT. Each dot represents the tumoral purity of the sample where >0.75 implies high tumoral purity and <0.25 represents low purity. **B)** Tumoral ploidy reported for chemoresistant (left, orange) and chemosensitive (right, blue) NS-TGCT. Each dot represents the tumoral ploidy of the sample where >1 implies high tumoral ploidy and <1 represents low ploidy.

**S3**.

Comparison of amp/del per segment over resistant (up) and sensitive (down) from GISTIC analysis **A) & B)** Amp/Del events (red/blue, respectively) in resistant samples. Left bar represents a total chromosome array, each line indicated are labeled genomic segments and its longitud represents the frequency of each event. G-Score corresponds to the peaks with higher incidences, estimated for gain or loss in those genomic segments. Scheme is the same for **C) & D)**. Broadly, the GISTIC plot reveals that platinum-sensitive samples have more gain and loss sites than resistant and copies are lost in different segments more frequently than are lossed.

**S4**.

Visualization of aCGH in Chr2 region in two samples to compare focal-CNVs inl 2q11.1 segment **A)** A resistant sample (up) included in aCGH, shows the 2q11.1 segment without gains or losses (blue and red, respectively) **B)** A sensitive sample (down) included in aCGH shows the constant gains in the 2q11.1 segment in sensitive samples. Both **A) and B)** shows in right a zoom with the genes contained in the 2q11.1 region.

