## Supplementary figures and images for "Genomic profile in TGCT Mexican patients reveals a potential biomarker of sensitivity to platinum-based therapy"

### Supplemental figures 1-4

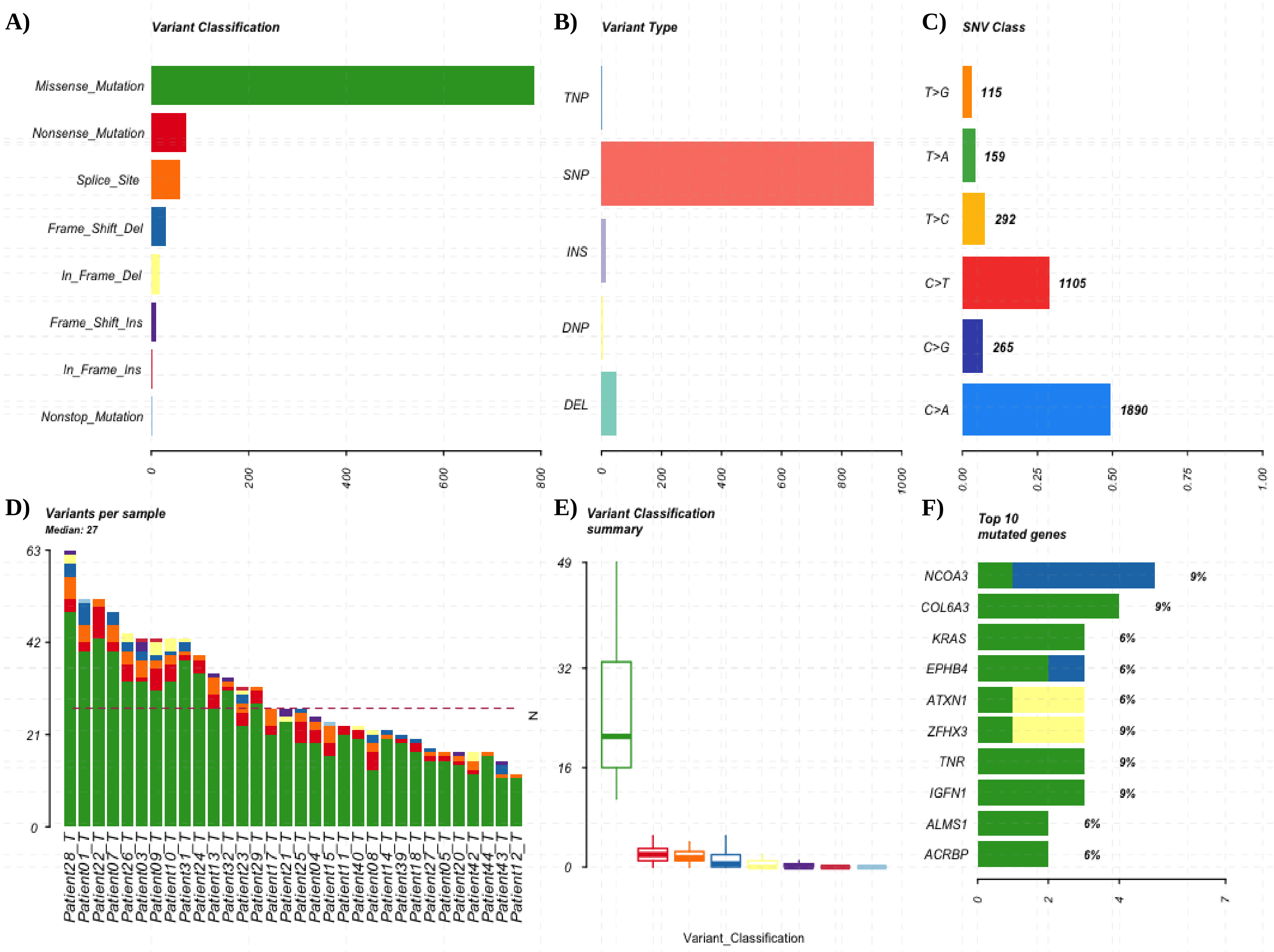


**Supplementary figure 1**

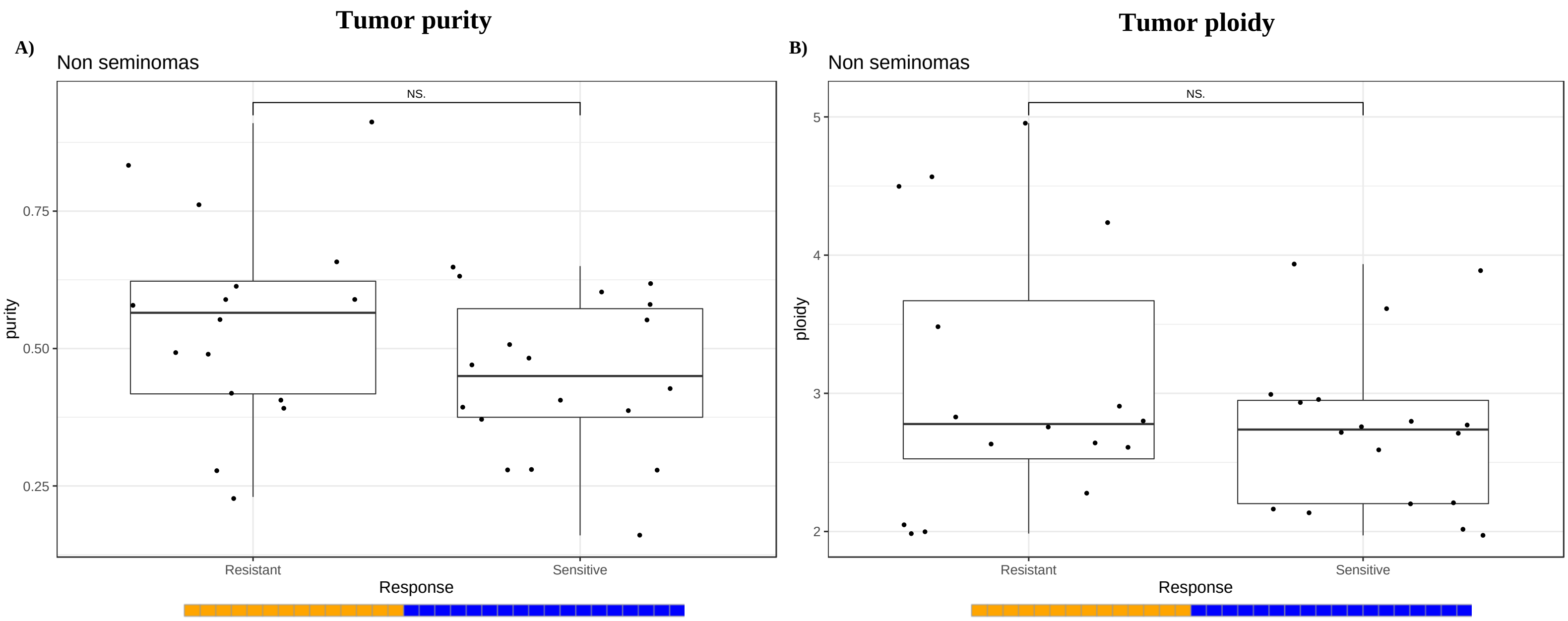

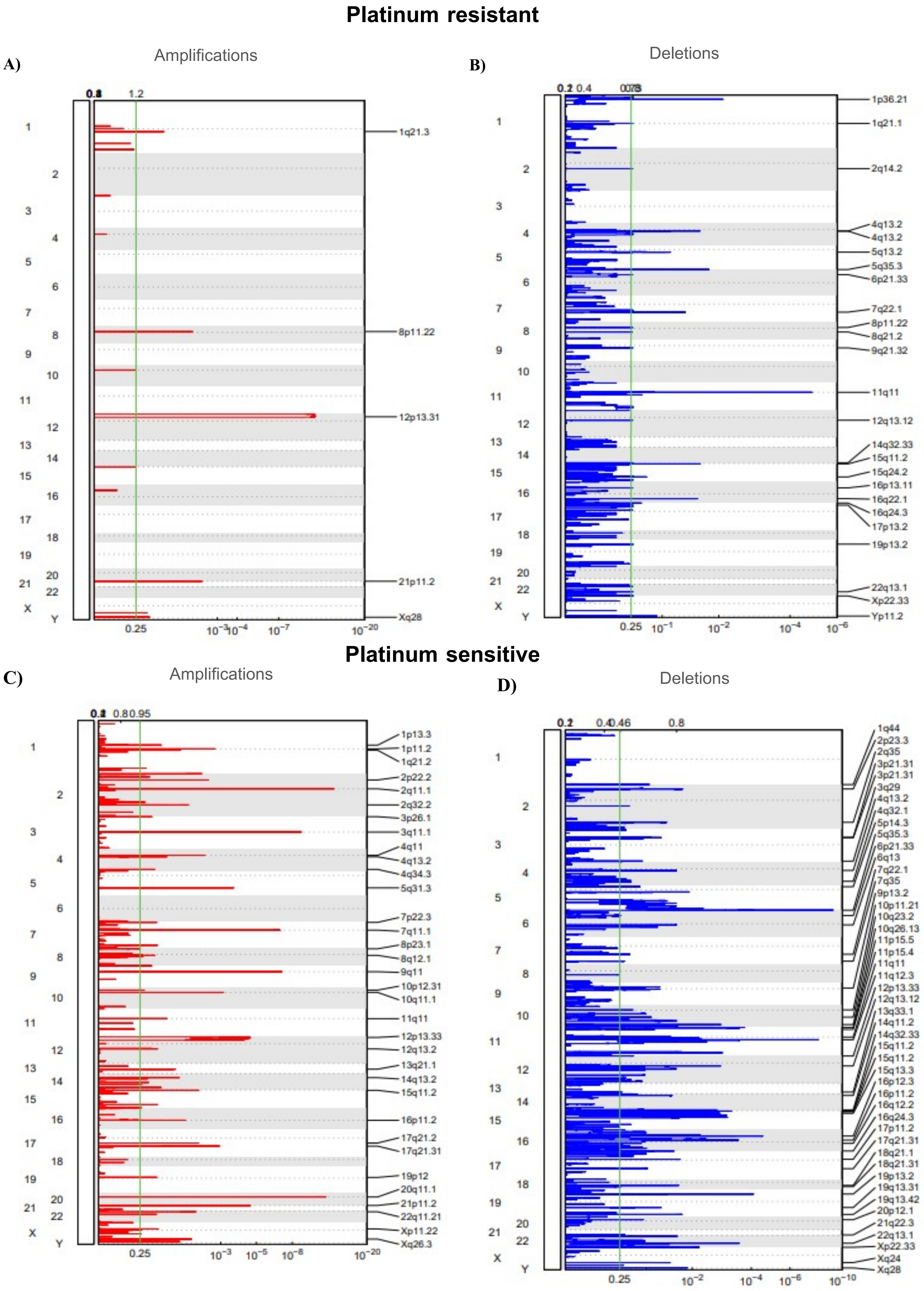

**A)** Resistant 2q11.1

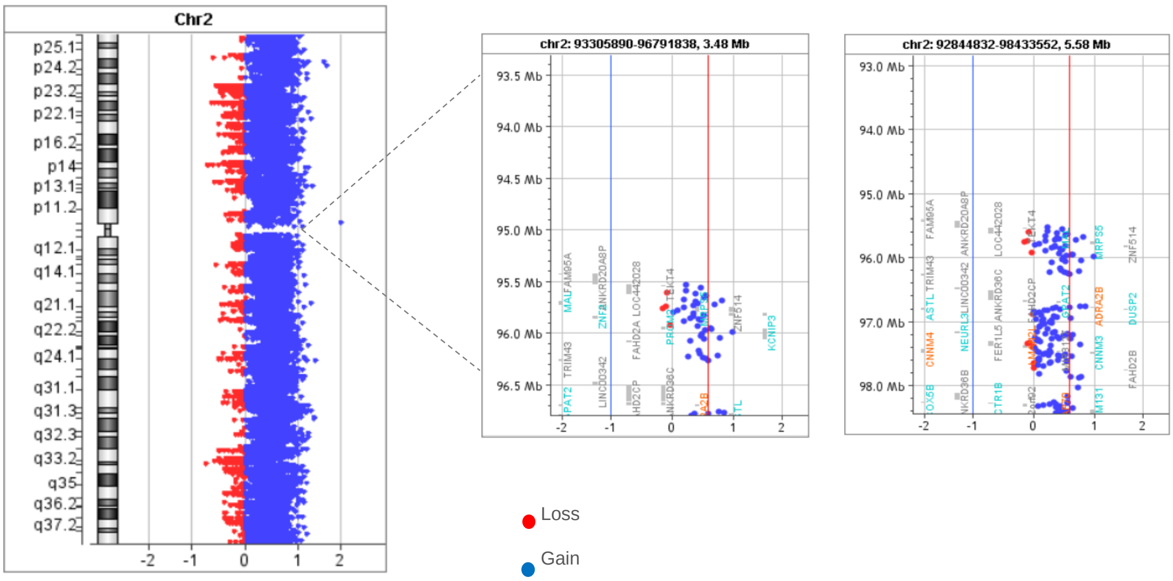


**B)** Sensitive 2q11.1

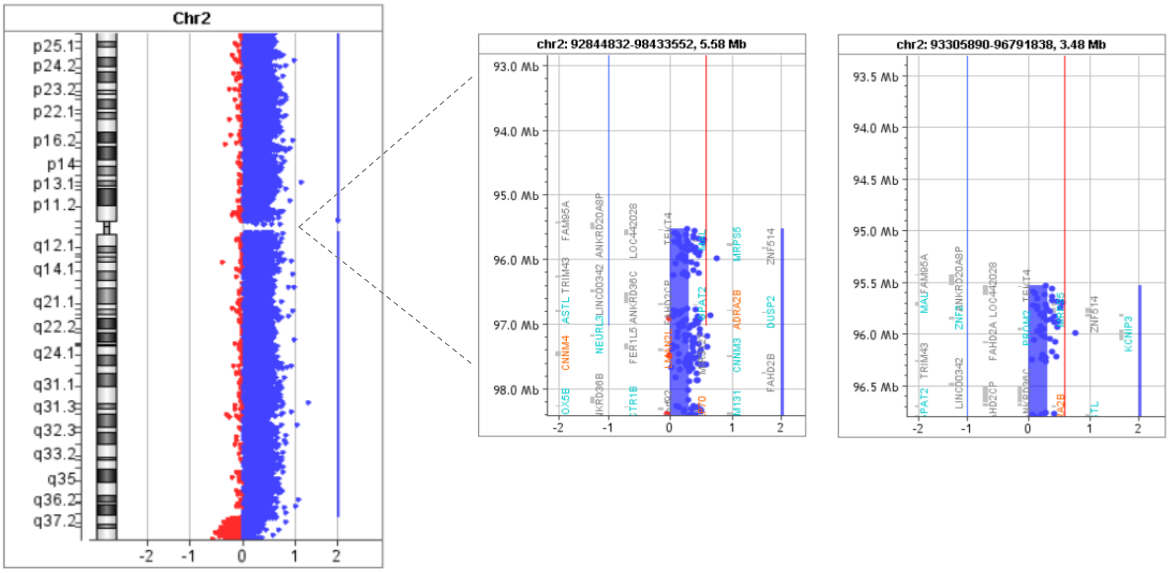


**Supplementary figure 4**
